# Trends in Prevalence, Risk-Factors Controls and Medications in ASCVD Among US Adults, 1999-2018

**DOI:** 10.1101/2023.02.24.23286436

**Authors:** Xiaowen Zhang, Jianzhou Chen, Zheng Chen, Chen Han, Lina Kang, Lian Wang, Kun Wang, Wei Xu, Biao Xu, Xinlin Zhang

**Author notes:** **Address for correspondence:** Xinlin Zhang, Department of Cardiology, Affiliated Drum Tower Hospital, Nanjing University School of Medicine, 321 Zhongshan Road, 210008 Nanjing, China. Xiaowen Zhang, Jianzhou Chen and Zheng Chen contributed equally to this work.

## Abstract

**Objective:** To characterize trends in prevalence of ASCVD, risk-factor control and medication use among adults with atherosclerotic cardiovascular disease (ASCVD), with frequent update of relevant guidelines.

**Patients and Methods:** We conducted a cross-sectional analysis of data from 55,081 adults in the National Health and Nutrition Examination Surveys (NHANES) 1999-2018.

**Results:** The age-standardized prevalence of ASCVD did not change significantly from 1999-2002 (7.9%, CI 7.1%-8.7%) to 2015-2018 (7.5%, CI 6.8%-8.3%) (*P* for trend =0.18). Over 60.0% ASCVD participants had very-high risk. The percentage with blood-pressure control (<130/80 mmHg) increased from 51.2% (CI, 41.0%-61.3%) in 1999-2002 to 57.2% (CI, 48.4%-65.6%) in 2011-2014, but then declined to 52.8% (CI, 44.4%-81.3%) in 2015-2018. From 1999-2002 to 2015-2018, the percentage with lipid control (non-high-density lipoprotein cholesterol <100 mg/dL) increased from 7.0% (CI, 3.5%-12.3%) to 26.4% (CI, 16.2%-38.9%), and with glycemic control (HbA1c <7.0%) decreased from 95.0% (CI, 90.2%-97.9%) to 84.0% (CI, 75.9%-90.3%). The percentage who achieved all 3 targets was 18.6% (CI, 8.2%-33.8%) in 2015-2018. After 2014, the percentages with blood-pressure, lipid, and glycemic control decreased in very-high-risk ASCVD, but increased in not-very-high-risk ASCVD. The percentage of ASCVD participants who used statins increased from 1999-2002 to 2011-2014, but then leveled off. The percentage who used blood-pressure-lowering drugs remained largely constant, and who used glucose-lowering drugs increased.

**Conclusions:** The prevalence of ASCVD generally remained stable, with over 60.0% had very-high risk. Blood-pressure, lipid, and glycemic control decreased in very-high-risk ASCVD but increased in not-very-high-risk ASCVD after 2014.

## Introduction

Decades of primary and secondary preventive efforts lead to substantial declines in cardiovascular (CV) mortality since 1980s to 2010s in the US. Atherosclerotic cardiovascular disease (ASCVD) remains the leading cause of morbidity and mortality ^1^. However, trends in prevalence of ASCVD, which includes coronary heart disease (CHD) and stroke, the top 2 major diseases that cause global CV mortality^1^ has not been documented. Since the last decade, the prevalence of dyslipidemia and smoking has decreased,^2, 3^ while prevalence of obesity and diabetes increased in US.^4, 5^ The temporal changes of risk factors in different directions might affect prevalence of ASCVD. Moreover, trends in risk-factor control and medication use might have changed since the last decade with frequent updates of relevant guidelines, such as the 2013 ACC/AHA guideline of blood cholesterol.^6^ which transitioned recommendations from “low-density lipoprotein cholesterol (LDL-C) target-based” to a “risk-based” approach, and the 2017 ACC/AHA guideline of high blood pressure,^7^ which lowered recommended goal of <130/80 mmHg for all adults taking blood pressure (BP)-lowering medications. The latest 2018 ACC/AHA cholesterol management guideline divides patients with ASCVD into those with very high risk and not-very-high risk.^8^ Patients with very-high risk have 3-fold greater risks for future ASCVD events.^9^ However, the proportion and trends of patients with very-high-risk ASCVD in US national population remain unclear.

This study aimed to provide nationally representative contemporary prevalence of ASCVD, describes trends in prevalence, risk-factor control and medication use, overall and by important subgroups (including ASCVD risk), among US adults from 1999 to 2018.

## Methods

### Data Source and Study Population

National Health and Nutrition Examination Surveys (NHANES) is a series of cross-sectional survey conducted every 2-year since 1999 by the National Center for Health Statistics of the Centers for Disease Control and Prevention. The survey examines a nationally representative sample of about 5,000 persons each year.^10^ The detailed study design and methods have been reported elsewhere.^11^ The NHANES program was approved by the National Center for Health Statistics Ethics Review Board, and all participants provided written consent.

Our study included adults aged 20 years or older who were nonpregnant from 10 consecutive cycles of NHANES between 1999-2000 and 2017-2018. ASCVD was self-reported with the questions “have you ever been told by a doctor or other health care professional that you had stroke, angina, myocardial infarction, or coronary heart disease?” Participants were defined to have ASCVD if their answer to any of the conditions was “yes”. This definition has been widely reported.^12-16^ Very-high-risk ASCVD was defined as multiple major ASCVD events or 1 major ASCVD event and multiple high-risk conditions.^17^ Major ASCVD events include recent angina or coronary heart disease (within 1 year), history of myocardial infarction, and history of stroke. High-risk conditions include age ≥65 years, familial hypercholesterolemia, diabetes mellitus, hypertension, chronic kidney disease, persistently elevated LDL-C (≥100 mg/dL) despite maximally tolerated statin therapy and ezetimibe, and history of congestive heart failure. We also performed a sensitivity analysis by not restricting angina or coronary heart disease to within 1 year.

### Risk Factors and Outcome Measures

Blood pressure (BP) measurements were obtained with a standardized protocol at a mobile examination center, and the average of all eligible BP readings were calculated. Hypertension was defined by a self-reported history of hypertension, current BP-lowering medication use, or with mean systolic BP ≥140 mmHg and/or diastolic BP ≥90 mmHg at examination.

Blood samples were collected at the time of the participant’s examination, and laboratory tests on these samples were performed using standard methods. Data extracted included total cholesterol (TC), high-density lipoprotein cholesterol (HDL-C), LDL-C, triglycerides, glycated hemoglobin (HbA1c), and fasting blood glucose (FBG). Non-HDL-C was calculated as the difference between TC and HDL-C. Diabetes was defined by a self-reported history of diabetes, currently taking glucose-lowering medications, or with FBG ≥126 mg/dL or HbA1c ≥6.5%. Hyperlipidemia was defined as self-reported history of hyperlipidemia or currently taking cholesterol-lowering medications.

We defined primary BP control as BP <130/80 mmHg,^7^ glycemic control as HbA1c <7.0% ^18^. The US guideline did not use target cholesterols, but it recommends an LDL-C threshold of 70 mg/dL or non-HDL-C threshold of 100 mg/dL to consider addition of nonstatins to statin therapy in ASCVD patients.^17^ Therefore, we defined primary lipid control as a non-HDL-C <100 mg/dL. We also set a BP target of <140/90 mmHg, non-HDL-C targets of <85 and 130 mg/dL, and LDL-C targets of <55, 70, and 100 mg/dL.^19^

### Medication Use

Uses of BP-lowering, glucose-lowering and lipid-lowering medications were assessed from medication review during a home visit. Medication names were self-reported and/or the drug containers were shown to the interviewers for verification. Medications were categorized into therapeutic classes with the Multum MediSource Lexicon classification system.^20^ BP-lowering medications were classified as angiotensin-converting enzyme (ACE) inhibitors or angiotensin receptor blockers (ARB), beta blockers, calcium channel blockers (CCB), diuretics, alpha blockers, centrally acting agents, direct vasodilators, renin inhibitors, and others. Glucose-lowering medications were classified as metformin, sulfonylureas, thiazolidinediones (TZD), dipeptidylpeptidase 4 (DPP-4) inhibitors, glucose like peptide-1 (GLP-1) receptor agonists, sodium-glucose co-transporter-2 (SGLT2) inhibitors, insulin, alpha-glucosidase inhibitors, meglitinides, amylin analogs, and others. The number and classes of BP-lowering and glucose-lowering medications were examined.

### Population Subgroups

Analyses were stratified by subgroups based on age (20-44, 45-64, or ≥65 years), sex (male or female), race and ethnicity (non-Hispanic white, non-Hispanic black, Mexican American, other Hispanic, or other/mixed), education (less than high school, high school graduate or general equivalency diploma, some college, or college graduate or higher), body mass index (BMI <25 [normal weight], 25–30 [overweight], or ≥30 [obese]), smoking status (current smoker, former smoker, or never smoked), and family income (income-to-poverty ratio <1.0, 1.0-2.99, 3.0-4.99, or ≥5.0). Race and ethnicity were self-reported, and non-Hispanic Asian was combined with “other race” in the analysis since it became a separate category from other races only after 2011. BMI was calculated as the weight in kilograms divided by height in meters squared. Income to poverty ratio was used to define socioeconomic status of the family, accounting for household size.

### Statistical Analyses

All analyses accounted for the NHANES complex survey design to ensure nationally representative estimates in accordance with recommended analytical guidelines ^21^. To produce estimates with greater precision and sampling error, we combined two adjacent 2-year cycles of the continuous NHANES into 4-year intervals.^21^ Categorical variables are reported as proportions (95% CI) and continuous variables as mean (95% CI). The proportions or means of ASCVD, CV risk factors and risk-factor control were calculated separately using data from each of the two 2-year cycles. Stratified analyses with the aforementioned subgroups were conducted. Analyses were age-standardized to the 2000 US Census population with the age categories 20-44 years, 45-64 years, and 65 years or older.^22^

Taylor series (linearization) method was used to estimate standard errors, and the Korn and Graubard method to estimate the 95% CI for prevalence.^23^ Trends over time were analyzed with linear regression models for age- and sex-adjusted means and with logistic regression models for proportions by adding the survey cycle as a continuous variable.^24^ In addition, Joinpoint regression analyses were used to determine trends in log-transformed age-standardized prevalence between time periods, allowing 1 joinpoint.^25^ The overall trend was initially estimated with no joinpoint and then a Monte Carlo permutation test was used to test significance of improvement in model fit by adding jointpoints. Factors associated with ASCVD and achieving risk-factor control were assessed using multivariate logistic regression models. To increase the statistical power, proportion of risk-factor control in subgroups were analyzed by combing all 10 year-cycles. Statistical analyses were performed with Stata software, version 16.0 (StataCorp) and Joinpoint Regression Program, version 4.9.1.0 (National Cancer Institute), and data management were conducted with R statistical computing software, version 3.5.2 (R Foundation). All reported *P* values were based on 2-sided tests with *P* <0.05 considered statistically significant.

## Results

### Participant Characteristics

A total of 55,081 adults were included in the analysis of ASCVD prevalence. 5,786 adults with ASCVD were included in the analyses of risk-factor control and medication use. From 1999 to 2018, the age, sex, and risk distribution of participants with ASCVD remained stable. The percentage of ASCVD participants of other/mixed ethnicity, with higher degree and BMI increased (Table S1).

### Trends in Prevalence of ASCVD

In 2015-2018, the prevalence of ASCVD in US adults was 7.5% (95% CI, 6.8% to 8.3%). About 60.0% of ASCVD participants had very-high risk. The prevalence of CHD and stroke was 5.7% (95% CI, 5.0% to 6.5%) and 2.7% (95% CI, 2.4% to 3.1%) respectively. The overall prevalence of ASCVD from 1999 to 2018 was significantly higher among older than young participants, men than women, White Americans than Mexican Americans and other Hispanic Americans, adults with a lower than higher education level, adults with a higher than lower BMI, former and current smoker than people never smoked, and people with a lower than higher family income, all after multivariable adjustment (Table 1 and Table S2).

**Table 1.** Trends in prevalence of ASCVD among US adults, 1999-2018.

|  | Adults with ASCVD, % 95 CI<sup>a</sup> |  |  |  |  |  |
| --- | --- | --- | --- | --- | --- | --- |
| Characteristics | 1999-2002 | 2003-2006 | 2007-2010 | 2011-2014 | 2015-2018 | P for trend<sup>b</sup> |
| No. with ASCVD<sup>c</sup> | 1090 | 1130 | 1300 | 1069 | 1197 |  |
| Overall prevalence | 7.9 (7.1, 8.7) | 8.4 (7.5, 9.3) | 7.7 (7.0, 8.5) | 7.6 (7.0, 8.3) | 7.5 (6.8, 8.3) | 0.18 |
| Age group, yr |  |  |  |  |  |  |
| 20-44 | 1.5 (1.2, 2.0) | 1.5 (1.0, 2.0) | 1.4 (1.0, 1.9) | 1.7 (1.2, 2.3) | 1.4 (1.0, 1.8) | 0.532 |
| 45-64 | 8.6 (7.1, 10.4) | 8.4 (6.8, 10.1) | 8.2 (7.1, 9.4) | 7.2 (6.3, 8.1) | 8.8 (7.4, 10.3) | 0.648 |
| ≥65 | 24.7 (22.0, 27.5) | 28.4 (25.6, 31.3) | 25.1 (22.2, 28.1) | 25.5 (23.6, 27.5) | 23.0 (20.7, 25.5) | 0.364 |
| Sex |  |  |  |  |  |  |
| Men | 9.5 (8.3, 10.8) | 9.4 (8.3, 10.6) | 9.4 (8.5, 10.3) | 9.1 (8.2, 10.1) | 9.1 (8.2, 10.2) | 0.026 |
| Women | 6.5 (5.6, 7.4) | 7.6 (6.6, 8.8) | 6.4 (5.5, 7.3) | 6.4 (5.5, 7.4) | 6.1 (5.2, 7.1) | 0.373 |
| Race and ethnicity |  |  |  |  |  |  |
| Non-Hispanic white | 8.0 (7.1, 9.1) | 8.4 (7.5, 9.5) | 7.6 (6.7, 8.7) | 7.7 (6.8, 8.5) | 7.4 (6.5, 8.4) | 0.088 |
| Non-Hispanic black | 9.0 (7.7, 10.5) | 9.0 (7.8, 10.4) | 8.6 (7.5, 9.9) | 7.8 (6.8, 9.0) | 8.3 (7.2, 9.6) | 0.138 |
| Mexican American | 6.3 (5.3, 7.4) | 7.0 (5.9, 8.3) | 7.2 (5.9, 8.6) | 6.6 (5.3, 8.2) | 5.3 (4.3, 6.6) | 0.836 |
| Other Hispanic | 4.9 (3.1, 7.3) | 3.6 (1.7, 6.5) | 6.4 (5.0, 8.0) | 6.6 (5.1, 8.3) | 7.2 (5.6, 9.2) | 0.084 |
| Other/mixed | 7.7 (4.4, 12.4) | 10.2 (6.2, 15.5) | 7.7 (5.6, 10.4) | 7.2 (5.7, 8.9) | 8.8 (6.5, 11.6) | 0.921 |
| Education level |  |  |  |  |  |  |
| Less than high school | 10.8 (9.3, 12.4) | 11.4 (9.6, 13.3) | 10.2 (9.1, 11.5) | 9.8 (8.5, 11.2) | 9.4 (7.5, 11.5) | 0.057 |
| High school graduate or GED | 8.3 (7.0, 9.7) | 7.9 (6.4, 9.5) | 8.7 (9.1, 7.3) | 9.5 (8.0, 11.2) | 8.8 (7.4, 10.5) | 0.169 |
| Some college | 7.4 (6.2, 8.8) | 9.1 (8.0, 10.3) | 7.1 (6.2, 8.2) | 6.9 (5.8, 8.2) | 8.1 (7.0, 9.3) | 0.693 |
| College graduate or higher | 5.3 (4.1, 6.7) | 6.0 (4.9, 7.2) | 5.4 (4.5, 6.5) | 5.6 (4.7, 6.6) | 5.2 (4.2, 6.3) | 0.492 |
| BMI range |  |  |  |  |  |  |
| <25 | 5.8 (4.9, 6.9) | 6.4 (5.4, 7.6) | 6.1 (5.1, 7.2) | 6.4 (5.4, 7.5) | 5.9 (4.9, 7.0) | 0.845 |
| 25-30 | 7.5 (6.5, 8.7) | 8.0 (7.1, 9.1) | 7.1 (6.3, 8.0) | 6.8 (6.0, 7.7) | 6.5 (5.5, 7.6) | 0.062 |
| ≥30 | 9.7 (8.4, 11.2) | 10.1 (8.7, 11.5) | 9.4 (8.3, 10.7) | 9.0 (8.0, 10.0) | 9.0 (8.0, 10.1) | 0.057 |
| Smoking status |  |  |  |  |  |  |
| Never smoked | 6.3 (5.5, 7.2) | 6.7 (5.8, 7.7) | 6.1 (5.4, 6.9) | 5.8 (5.1, 6.6) | 5.4 (4.8, 6.1) | 0.044 |
| Former smoker | 9.6 (8.3, 11.0) | 9.1 (7.8, 10.6) | 9.5 (8.2, 10.9) | 7.8 (6.7, 8.9) | 9.0 (7.3, 11.0) | 0.211 |
| Current smoker | 9.3 (7.4, 11.5) | 9.7 (7.9, 11.8) | 10.1 (8.7, 11.7) | 12.2 (9.7, 15.1) | 11.6 (9.7, 13.7) | 0.039 |
| Income to poverty ratio |  |  |  |  |  |  |
| <1.0 | 10.8 (9.0, 12.8) | 12.4 (10.6, 14.5) | 11.9 (10.4, 13.5) | 11.0 (9.4, 12.7) | 10.4 (8.9, 12.1) | 0.389 |
| 1.0-2.99 | 8.8 (7.6, 10.1) | 10.0 (8.8, 11.2) | 8.7 (7.8, 9.7) | 9.7 (8.4, 11.1) | 9.3 (7.9, 10.8) | 0.8 |
| 3.0-4.99 | 6.9 (5.6, 8.4) | 6.5 (4.9, 8.4) | 6.2 (5.1, 7.4) | 6.5 (5.4, 7.7) | 6.1 (4.8, 7.6) | 0.161 |
| ≥5.0 | 6.8 (5.3, 8.6) | 5.9 (4.5, 7.5) | 6.2 (5.0, 7.6) | 4.2 (3.3, 5.3) | 5.4 (4.2, 6.7) | 0.207 |

Abbreviations: ASCVD, atherosclerotic cardiovascular disease; BMI, body mass index; GED, general equivalency diploma.
<sup>a</sup>Estimates were standardized to the 2000 US Census using the age categories 20–44 years, 45–64 years, and 65 years or older. <sup>b</sup>P values were obtained from the Jointpoint Regression program. <sup>c</sup>Unweighted number of adults with ASCVD.

The prevalence of ASCVD did not change significantly from 1999-2002 to 2015-2018 (*P* for trend =0.18), as were prevalence of very-high risk and not-very-high risk ASCVD (*P* for trend =0.44 and 0.15 respectively) and prevalence of CHD and stroke (*P* for trend =0.17 and 0.81 respectively) (Figure 1). When CHD event was not restricted to within 1 year, 8.7% of adults with ASCVD had multiple ASCVD events, 61.4% had 1 ASCVD event and multiple high-risk conditions, and 9.3% had 1 ASCVD event but no high-risk conditions in 2015-2018 (Figure S1). The percentage of ASCVD participants with multiple high-risk conditions or multiple events increased after 2007-2010 (Figure S1). No significant trends were found in prevalence of ASCVD across cycles stratified by age group, race and ethnicity, education level and family income (Table 1). Age-standardized prevalence of ASCVD did not change significantly for women, but a decreasing trend was detected in men (*P* for trend =0.026). The prevalence of ASCVD decreased in participants who never smoked but increased in current smokers from 1999-2002 to 2015-2018 (*P* for trend =0.044 and 0.039 respectively).

**Figure 1.**
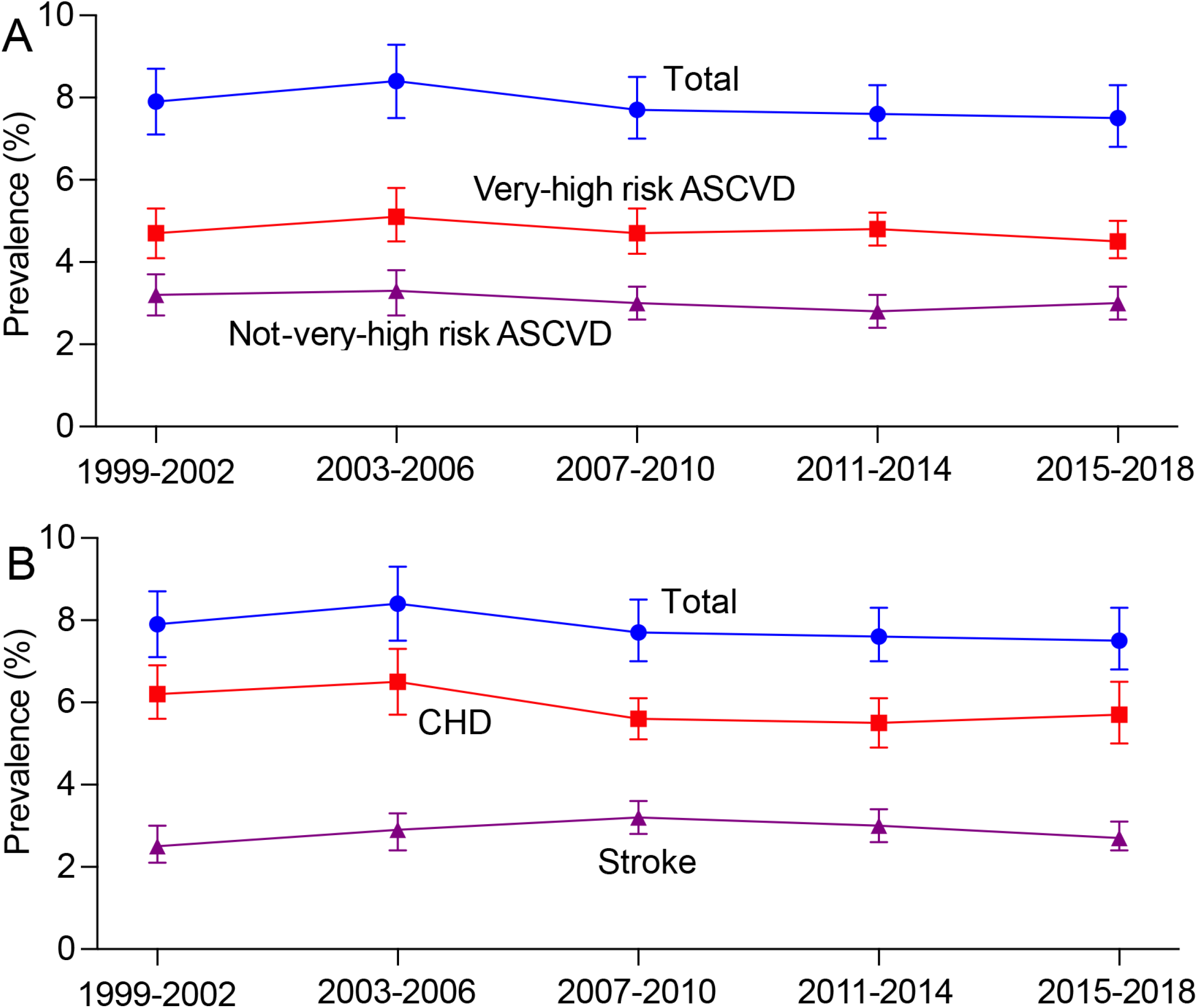
Trends in prevalence of ASCVD among US adults, 1999-2002 to 2015-2018. (A) Trends in prevalence of total ASCVD, very-high-risk ASCVD, and not-very-high-risk ASCVD; (B) Trends in prevalence of total ASCVD, coronary heart disease (CHD), and stroke. Abbreviation: ASCVD, atherosclerotic cardiovascular disease.

### Trends in Risk-factor Control

The age-adjusted mean HbA1c increased from 5.6% in 1999-2002 to 6.2% in 2015-2018, as were FBG, waist circumference and BMI (all *P* for trend <0.001). The age-adjusted mean non-HDL-C decreased from 160.1 mg/dL in 1999-2002 to 128.6 mg/dL in 2015-2018 (*P* for trend <0.001). Similar trends of decrement were found for TC, LDL-C and triglycerides. There were no statistically significant linear trends in overall BP across year cycles, but mean BP increased after 2010 (Table S3).

The percentage of adults who achieved BP control of <130/80 mmHg increased from 51.2% (95% CI 41.0% to 61.3%) in 1999-2002 to 57.2% (95% CI 48.4% to 65.6%) in 2011-2014, but then declined to 52.8% (95% CI 44.4% to 81.3%) in 2015-2018 (Figure 2). The percentage of adults who achieved lipid control (non-HDL-C <100 mg/dL) increased from 7.0% (95% CI 3.5% to 12.3%) in 1999-2002 to 26.4% (95% CI 16.2% to 38.9%) in 2015-2018 (*P* for trend <0.001). The percentage of adults who achieved HbA1c <7.0% decreased from 95.0% (95% CI 90.2% to 97.9%) to 84.0% (95% CI 75.9% to 90.3%) (*P* for trend <0.001). The percentage of participants who achieved all 3 risk-factor control (HbA1c <7.0%, BP <130/80 mmHg and non-HDL-C <100 mg/dL) increased from 4.8% (95% CI 1.8% to 10.3%) to 18.6% (95% CI 8.2% to 33.8%) (*P* for trend <0.001) (Figure 2, Table S4).

**Figure 2.**
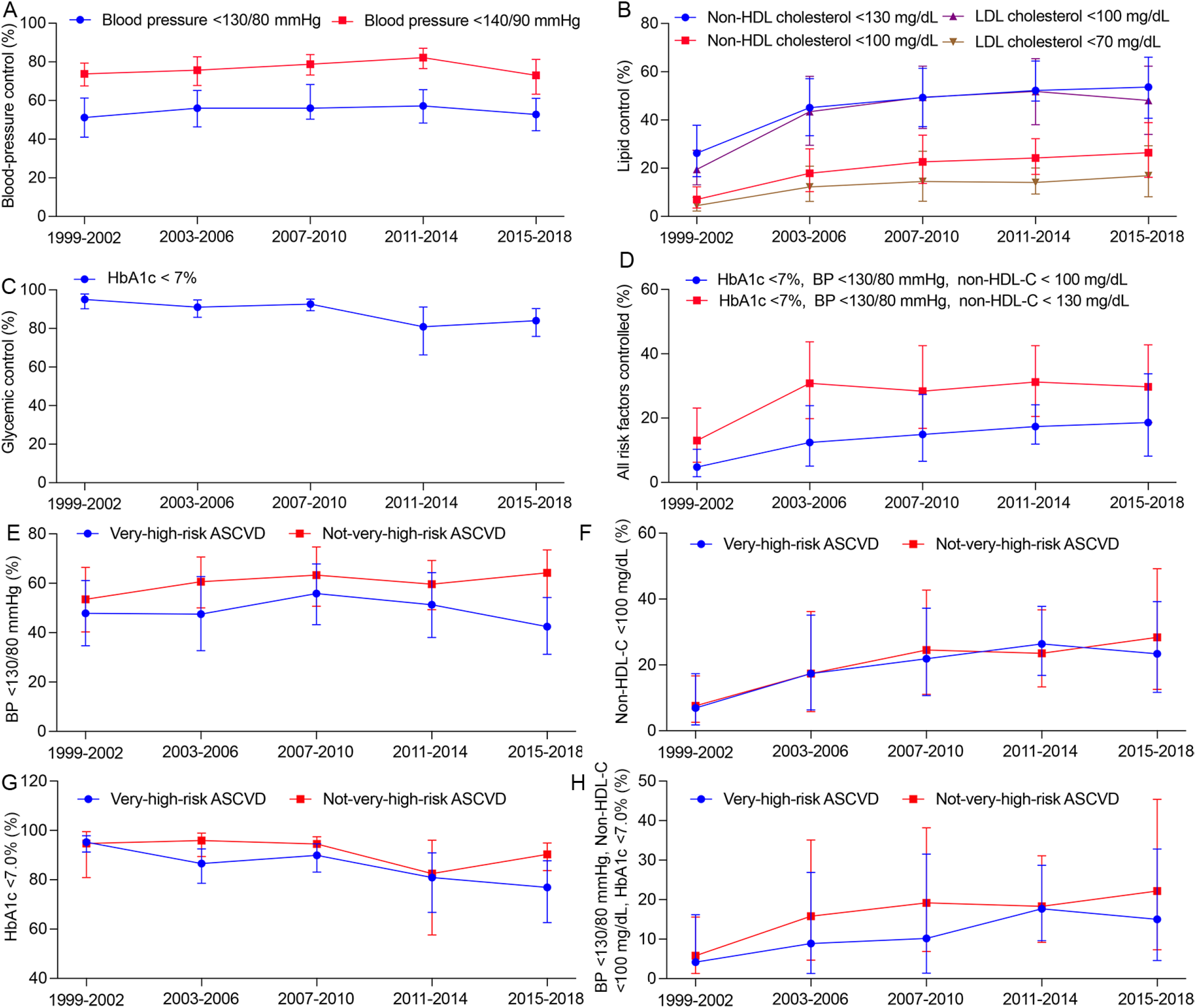
Trends in risk-factor control among US adults with ASCVD, 1999-2002 to 2015-2018. (A-D) Trends in prevalence of primary and secondary BP, lipid, and glycemic control among US adults with ASCVD, 1999-2002 to 2015-2018. (E-H) Trends in prevalence of primary BP, lipid, and glycemic control among US adults with ASCVD stratified by ASCVD risk, 1999-2002 to 2015-2018. Abbreviations: ASCVD, atherosclerotic cardiovascular disease; HbA1c, glycated hemoglobin (HbA1c); LDL-C, low-density lipoprotein cholesterol; Non-HDL-C, non-high-density lipoprotein cholesterol.

Subgroup estimates for risk-factor control are shown in Table 2, with data from the entire study period combined. Compared with younger adults, the older were less likely to achieve primary BP target (OR, 0.47 [95% CI, 0.32 to 0.68]). Women were more likely to achieve glycemic target than men (OR, 2.0 [95% CI, 1.41 to 2.82]). Compared with non-Hispanic White, non-Hispanic Black were less likely to achieve primary BP target (OR, 0.63 [95% CI, 0.53 to 0.77]), but more likely to achieve primary lipid target (OR, 1.61 [95% CI, 1.17 to 2.22]). Compared with non-Hispanic White, all other races were less likely to achieve glycemic target. Adults with higher BMI, lower education level and lower income were less likely to achieve ≥2 of the 3 risk-factor targets (Table 2 and Table S5).

**Table 2.** Prevalence of achieving risk-factor control among US adults with ASCVD, 1999-2018.

| Characteristics | Adults with ASCVD, % (95% CI)<sup>a</sup> |  |  |  |  |  |  |  |  |
| --- | --- | --- | --- | --- | --- | --- | --- | --- | --- |
|  | BP <130/80 mmHg | BP <140/90 mmHg | Non-HDL-C <100 mg/dL | Non-HDL-C <130 mg/dL | LDL-C <70 mg/dL | LDL-C <100 mg/dL | HbA1c <7.0% | HbA1c <7%, BP <130/80 mmHg, non-HDL-C < 100 mg/dL | HbA1c <7%, BP <130/80 mmHg, non-HDL-C < 130 mg/dL |
| No.<sup>b</sup> | 5100 | 5100 | 5011 | 5011 | 2315 | 2315 | 5106 | 5343 | 5343 |
| Overall prevalence | 55.4 (51.4, 59.3) | 76.9 (74.0, 79.7) | 20.3 (16.6, 24.5) | 46.1 (40.8, 51.4) | 13.0 (9.9, 16.6) | 43.4 (37.6, 49.4) | 88.4 (84.9, 91.4) | 14.1 (10.4, 18.4) | 27.2 (22.3, 32.7) |
| Age group, yr |  |  |  |  |  |  |  |  |  |
| 20-44 | 62.6 (55.0, 70.0) | 82.8 (77.2, 87.4) | 20.3 (13.3, 28.9) | 44.1 (34.3, 54.2) | 10.0 (4.5, 18.7) | 37.9 (27.4, 49.2) | 91.8 (81.9, 96.5) | 17.6 (10.8, 26.2) | 31.6 (22.7, 41.7) |
| 45-64 | 50.0 (46.4, 53.4) | 75.2 (72.2, 78.0) | 15.9 (12.2, 20.2) | 43.3 (38.6, 48.2) | 12.8 (9.1, 17.4) | 44.4 (39.2, 49.8) | 83.5 (79.5, 87.1) | 9.2 (6.4, 12.7) | 22.2 (17.8, 27.2) |
| ≥65 | 43.9 (41.6, 46.1) | 63.2 (60.9, 65.5) | 28.2 (24.7, 31.9) | 56.7 (53.3, 60.1) | 21.7 (19.0, 24.7) | 57.7 (54.5, 60.9) | 87.3 (84.6, 89.7) | 12.4 (10.1, 15.1) | 23.3 (20.3, 26.5) |
| Sex |  |  |  |  |  |  |  |  |  |
| Men | 51.2 (45.8, 56.5) | 77.8 (74.0, 81.4) | 18.7 (13.7, 24.7) | 38.9 (32.1, 46.0) | 13.5 (9.5, 18.5) | 37.5 (30.5, 45.0) | 85.0 (78.1, 90.3) | 12.5 (7.6, 18.9) | 20.3 (14.6, 27.1) |
| Women | 59.0 (54.4, 63.4) | 75.5 (71.6, 79.1) | 20.5 (14.9, 27.0) | 50.3 (43.1, 57.4) | 11.6 (7.1, 17.5) | 45.9 (38.3, 53.6) | 91.6 (88.8, 93.9) | 14.5 (9.5, 20.9) | 32.1 (25.0, 40.0) |
| Race and ethnicity |  |  |  |  |  |  |  |  |  |
| Non-Hispanic white | 57.8 (52.2, 63.4) | 81.1 (77.7, 84.3) | 18.8 (13.8, 24.7) | 44.0 (36.7, 51.6) | 13.5 (9.3, 18.8) | 42.4 (34.5, 50.7) | 90.7 (85.9, 94.2) | 14.2 (9.3, 20.3) | 27.7 (20.9, 35.3) |
| Non-Hispanic black | 42.0 (36.5, 47.7) | 59.8 (53.8, 65.6) | 27.3 (18.6, 37.5) | 48.5 (38.7, 58.4) | 14.8 (8.5, 23.2) | 44.4 (34.2, 54.9) | 83.4 (75.1, 89.9) | 15.3 (8.2, 25.2) | 23.8 (15.2, 34.3) |
| Mexican American | 57.7 (49.6, 65.6) | 74.9 (67.0, 81.8) | 21.9 (12.6, 34.0) | 49.7 (36.2, 63.3) | 10.6 (6.2, 16.7) | 45.4 (32.8, 58.5) | 77.8 (65.2, 87.4) | 13.1 (5.4, 25.3) | 26.6 (15.5, 40.2) |
| Other Hispanic | 59.9 (47.4, 71.6) | 72.7 (58.5, 84.2) | 20.1 (9.3, 35.4) | 42.9 (28.0, 58.9) | 10.4 (4.7, 19.3) | 41.7 (26.5, 58.3) | 83.8 (70.4, 92.8) | 13.6 (4.2, 30.0) | 25.9 (13.2, 42.4) |
| Other/mixed | 51.8 (38.5, 64.9) | 81.5 (72.6, 88.4) | 19.5 (8.8, 35.0) | 61.9 (41.9, 79.3) | 8.1 (3.3, 16.1) | 46.2 (27.3, 65.8) | 89.8 (83.2, 94.5) | 5.7 (1.5, 14.5) | 22.7 (8.0, 44.9) |
| Education level |  |  |  |  |  |  |  |  |  |
| Less than high school | 53.7 (47.2, 60.1) | 73.7 (69.1, 78.0) | 17.8 (11.6, 25.5) | 45.2 (36.6, 54.0) | 9.9 (6.8, 13.8) | 38.7 (30.8, 47.1) | 89.0 (85.2, 92.2) | 11.2 (5.9, 18.9) | 27.6 (19.3, 37.2) |
| High school graduate or GED | 51.3 (44.2, 58.4) | 75.0 (68.4, 80.8) | 19.9 (13.6, 27.6) | 44.9 (35.9, 54.2) | 11.9 (7.1, 18.3) | 42.7 (33.3, 52.5) | 86.8 (79.6, 92.2) | 13.6 (7.3, 22.5) | 26.1 (17.7, 35.9) |
| Some college | 55.9 (49.2, 62.5) | 76.8 (71.8, 81.3) | 21.7 (14.8, 30.2) | 47.2 (37.9, 56.6) | 16.6 (11.2, 23.2) | 45.6 (36.0, 55.4) | 88.4 (81.4, 93.5) | 15.0 (8.0, 24.5) | 27.1 (19.5, 35.9) |
| College graduate or higher | 66.1 (57.2, 74.2) | 86.6 (82.5, 90.1) | 20.9 (11.1, 33.9) | 45.0 (29.1, 61.6) | 13.5 (4.7, 28.2) | 47.5 (30.3, 65.1) | 89.7 (81.5, 95.2) | 15.4 (6.3, 29.5) | 25.2 (13.2, 40.9) |
| BMI range |  |  |  |  |  |  |  |  |  |
| <25 | 64.8 (58.5, 70.7) | 79.4 (73.9, 84.1) | 40.4 (28.5, 53.3) | 65.7 (54.7, 75.6) | 21.3 (11.3, 34.6) | 58.9 (46.4, 70.5) | 95.8 (91.8, 98.2) | 29.5 (18.3, 43.0) | 46.1 (34.2, 58.3) |
| 25-30 | 57.9 (50.2, 65.3) | 78.7 (72.3, 84.1) | 17.1 (11.0, 24.8) | 50.3 (39.1, 61.5) | 8.4 (5.5, 12.1) | 45.0 (33.5, 57.0) | 93.8 (90.2, 96.4) | 12.5 (7.0, 20.0) | 30.9 (21.1, 42.2) |
| ≥30 | 50.7 (45.4, 55.9) | 75.5 (72.6, 79.1) | 15.5 (11.1, 20.9) | 37.3 (31.2, 43.8) | 13.2 (9.3, 18.0) | 37.5 (31.1, 44.2) | 81.9 (75.3, 87.4) | 9.5 (5.1, 15.9) | 18.6 (13.2, 25.1) |
| Smoking status |  |  |  |  |  |  |  |  |  |
| Never smoked | 54.3 (48.6, 60.0) | 77.5 (73.6, 81.0) | 20.2 (13.4, 28.5) | 44.4 (35.3, 53.8) | 9.9 (6.5, 14.4) | 42.8 (33.3, 52.7) | 87.3 (82.0, 91.5) | 14.3 (8.0, 22.9) | 27.2 (19.1, 36.6) |
| Former smoker | 53.7 (45.3, 61.9) | 75.6 (68.6, 81.8) | 22.9 (13.6, 34.6) | 46.9 (35.0, 59.0) | 13.2 (7.2, 21.6) | 46.1 (34.2, 58.4) | 82.9 (69.6, 92.1) | 17.6 (8.9, 29.9) | 25.9 (16.1, 37.8) |
| Current smoker | 57.7 (51.5, 63.7) | 77.2 (71.9, 81.9) | 19.0 (12.6, 26.7) | 46.0 (38.4, 53.8) | 14.4 (8.7, 21.8) | 42.2 (33.7, 51.0) | 92.7 (89.3, 95.3) | 11.9 (6.5, 19.3) | 28.0 (20.8, 36.1) |
| Income to poverty ratio |  |  |  |  |  |  |  |  |  |
| <1.0 | 56.1 (50.5, 61.6) | 71.4 (66.7, 75.8) | 20.1 (13.8, 27.7) | 44.1 (36.5, 51.9) | 13.0 (7.7, 20.0) | 37.8 (29.8, 46.4) | 89.4 (85.6, 92.5) | 13.8 (8.0, 21.9) | 25.3 (17.8, 34.1) |
| 1.0-2.99 | 51.1 (44.8, 57.3) | 76.5 (72.2, 80.5) | 21.0 (14.6, 28.6) | 41.0 (33.3, 48.9) | 15.7 (9.9, 23.2) | 39.2 (31.3, 47.5) | 87.5 (82.4, 91.6) | 14.0 (8.3, 21.7) | 21.7 (15.1, 29.5) |
| 3.0-4.99 | 61.3 (51.8, 70.2) | 81.6 (74.3, 87.6) | 24.5 (10.8, 43.4) | 56.1 (40.2, 71.2) | 9.3 (6.2, 13.4) | 45.9 (28.9, 63.7) | 91.2 (84.7, 95.6) | 20.7 (7.7, 40.3) | 36.2 (20.7, 54.2) |
| ≥5.0 | 53.9 (38.3, 69.1) | 84.3 (77.7, 89.5) | 11.7 (8.0, 16.2) | 40.5 (25.4, 57.0) | 8.4 (5.3, 12.6) | 47.9 (30.7, 65.5) | 83.7 (61.6, 95.9) | 7.0 (4.2, 10.8) | 23.7 (13.2, 37.2) |

Abbreviations: ASCVD, atherosclerotic cardiovascular disease; BMI, body mass index; BP, blood pressure; GED, general equivalency diploma; HbA1c, glycated hemoglobin (HbA1c); LDL-C, low-density lipoprotein cholesterol; Non-HDL-C, non-high-density lipoprotein cholesterol. <sup>a</sup>Prevalence estimates were standardized to the 2000 US Census using the age categories 20–44 years, 45–64 years, and 65 years or older. <sup>b</sup>Unweighted number of adults with ASCVD.

Compared with ASCVD adults who were not at very-high risks, very-high-risk adults were less likely to achieve primary BP target (OR, 0.78 [95% CI, 0.68 to 0.90]), glycemic target (OR, 0.64 [95% CI, 0.46 to 0.91]), and all 3 risk-factor targets (OR, 0.63 [95% CI, 0.42 to 0.94]) (Table S6), while primary lipid target was similarly achieved (OR, 1.06 [95% CI 0.79 to 1.43]). After 2014, the percentages of participants who achieved BP, lipid, and glycemic targets decreased in very-high-risk ASCVD, but increased in not-very-high-risk ASCVD (Figure 2, Table S7 and S8). The absolute values and trends of lipids, BP, glucose and other variables among ASCVD adults, overall and stratified by ASCVD risk are shown in Figure S2 and S3. Compared with adults with stroke, those with CHD were more likely to achieve BP and lipid targets, those with CHD and stroke were more likely to achieve lipid target (Table S9).

### Trends in Medication Use

The distribution of medication use among ASCVD adults is shown in Figure 3. The percentage of ASCVD participants who used statins increased from 40.1% in 1999-2002 to 63.4% in 2011-2014, but then leveled off (*P* for trend <0.001). The percentage of ezetimibe use increased to 7.0% in 2007-2010, but then declined to 1.6% in 2015-2018. The frequency of BP-lowering medication use remained largely constant between 72.6% and 78.8% from 1999-2002 to 2015-2018 (*P* for trend =0.06). The use of ACE inhibitors or ARB and beta-blockers increased over time, while frequencies of diuretics and CCB decreased. The frequency of any glucose-lowering medication use increased from 16.6% in 1999-2002 to 30.1% in 2015-2018 (*P* for trend <0.001). The use of metformin, SGLT2 inhibitors or GLP-1, and DPP-4 inhibitors increased, while the use of thiazolidinediones and sulfonylureas decreased since the last decade.

**Figure 3.**
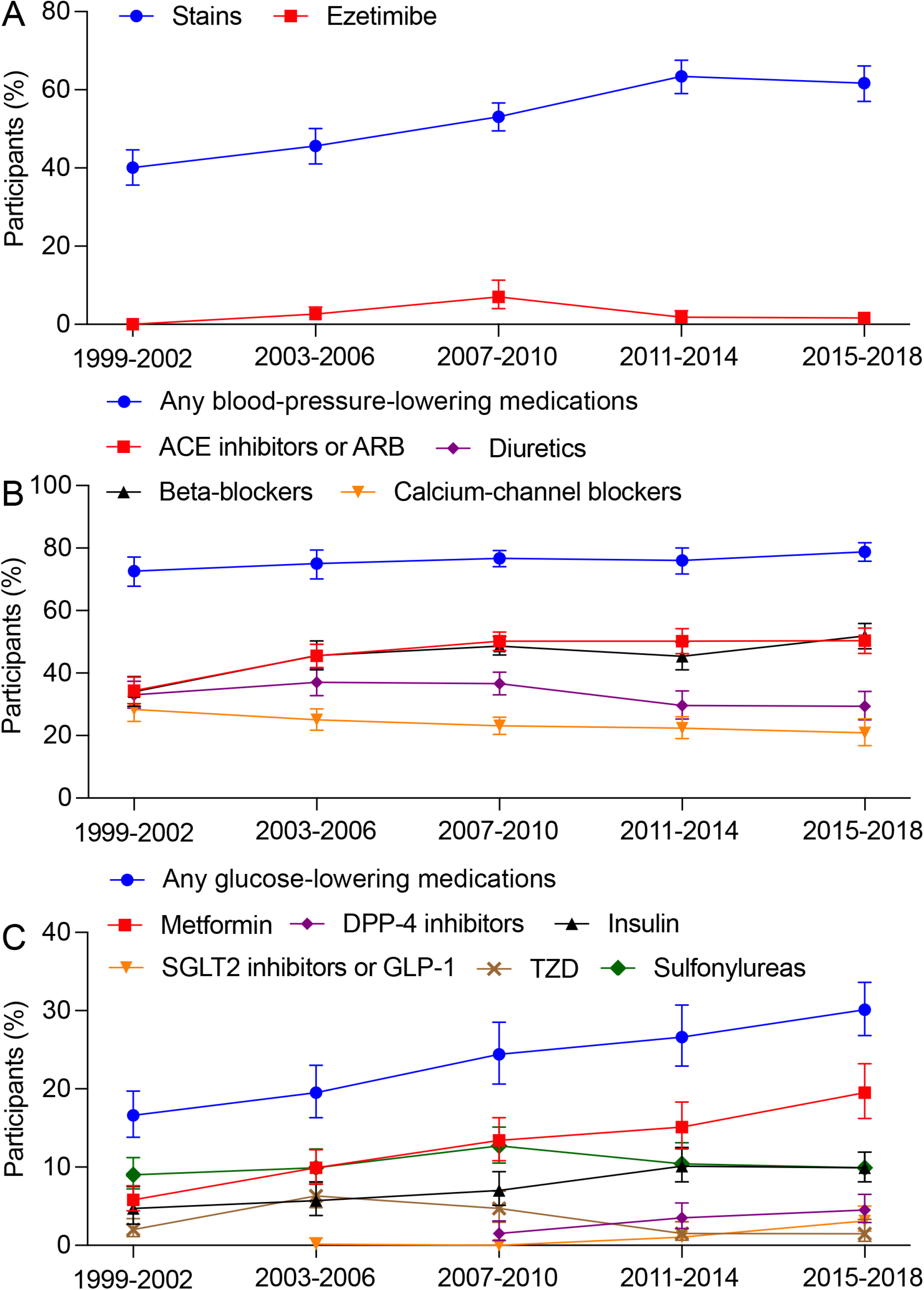
Trends in medications among US adults with ASCVD, 1999-2002 to 2015-2018. (A) Lipid-lowering medications. (B) Blood-pressure-lowering medications. (C) Glucose-lowering medications. Abbreviations: ACE, angiotensin-converting enzyme; ASCVD, atherosclerotic cardiovascular disease; ARB, angiotensin receptor blocker; CCB, calcium channel blocker; DPP-4, dipeptidylpeptidase 4; GLP-1, glucose like peptide-1 receptor agonists; SGLT2, sodium-glucose co-transporter-2; TZD, thiazolidinedione.

There was no significant linear trend in the use of 1, 2, or 3 or more classes of BP-lowering medications (Figure S4). The percentage of adults receiving combination BP-lowering therapy declined after 2007-2010. In 2015-2018, 50.5% of individuals were taking combination BP-lowering therapy. The proportion of ASCVD adults taking 1, 2, or 3 or more classes of glucose-lowering medications significantly increased across cycles (Figure S5). In 2015-2018, 13.5% of individuals were taking combination glucose-lowering medications.

In adults who did not achieve risk-factor control, those with younger age, women, non-Hispanic black, Mexican American, with lower BMI and income were less likely to receive statins, and adults with younger age and lower BMI were also less likely to receive BP-lowering treatment than their corresponding comparators (Table S10). Non-Hispanic Black were less likely to receive combination glucose-lowering therapy but were more likely to receive combination BP-lowering therapy when targets were not achieved (Table S10). Adults with stroke were less likely to receive statins and combination BP-lowering therapy than those with CHD (Table S11).

## Discussions

In this nationally representative serial cross-sectional study that included 55,081 adults, the age-standardized prevalence of ASCVD remained largely stable from 1999-2002 to 2015-2018 (ranges from 7.5-7.9%), with the percentage of coronary heart disease (CHD) doubles that of stroke (Central Figure). The stable prevalence of ASCVD during this period was also found in most subgroups. Blood-pressure and glycemic control declined while lipid control remained slight increment after 2010-2014, with less than 20% of ASCVD participants achieved all 3 risk-factor control in 2015-2018. 60.0% of ASCVD participants had very-high risk according to recent US guideline, and BP, lipid, and glycemic control decreased in very-high-risk ASCVD but increased in not-very-high-risk ASCVD after 2014. The percentage of ASCVD participants who used statins leveled off after 2014, and ezetimibe use decreased after 2010. The percentage who used any BP-lowering drugs remained largely constant, and who used any glucose-lowering drugs increased from 1999-2002 to 2015-2018. There were differences in percentage of participants achieving different risk-factor control and taking medications across age, racial or ethnic, gender, BMI, education and income groups.

**Central Figure.**
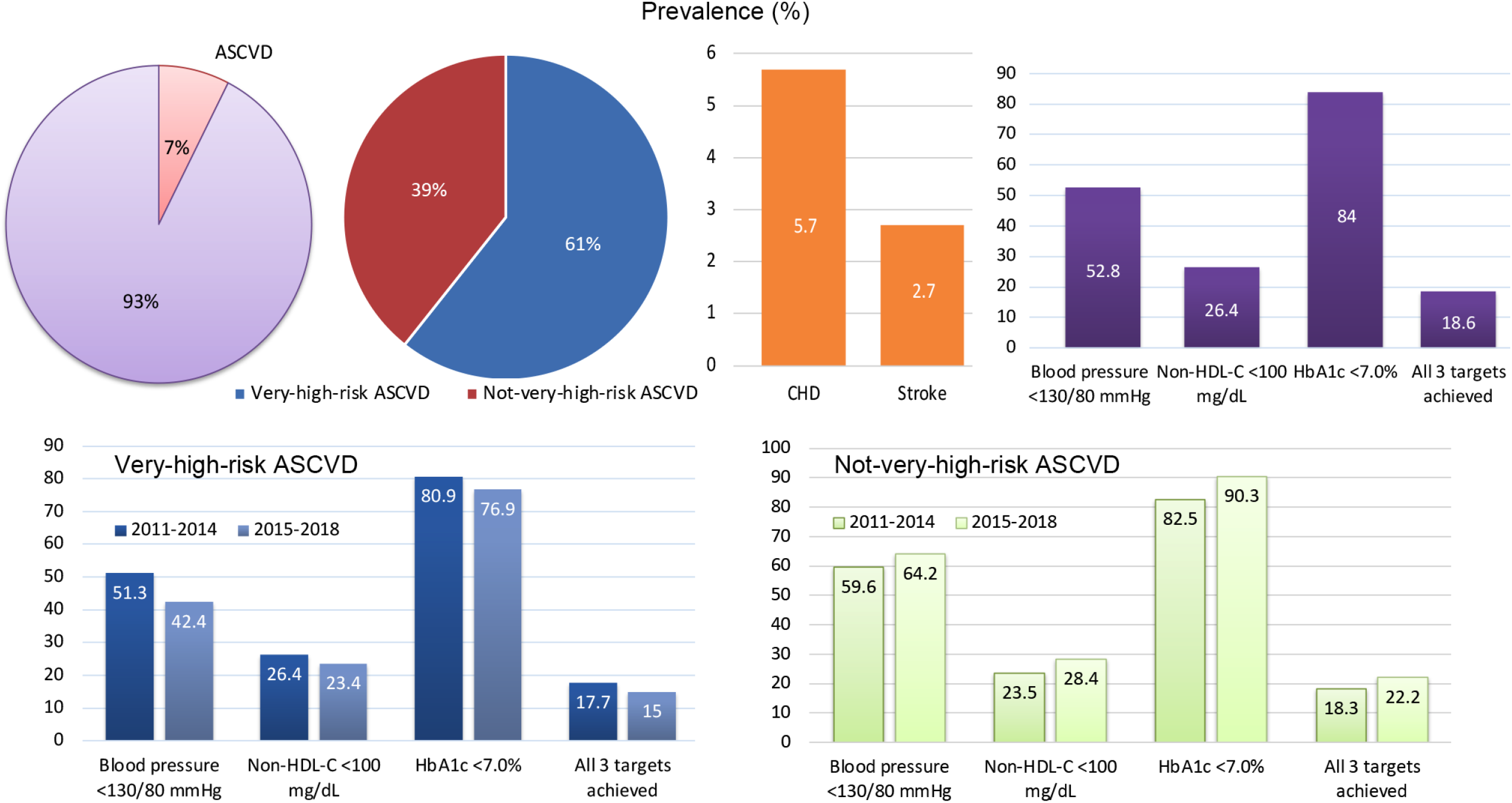
Prevalence of ASCVD and risk-factor control among US adults, 2011-2014 to 2015-2018. The age-standardized prevalence of ASCVD was 7.5% in 2015-2018, with the prevalence of coronary heart disease (CHD) doubles that of stroke. Among ASCVD adults, 61.0% had very-high risk. Blood-pressure, lipid, and glycemic control was 52.8%, 26.4% and 84% respectively, with 18.6% of ASCVD participants achieved all 3 risk-factor control in 2015-2018. The percentages of ASCVD adults with blood-pressure, lipid, and glycemic control decreased in very-high-risk ASCVD, but increased in not-very-high-risk ASCVD from 2011-2014 to 2015-2018.

We found that over 60% of ASCVD participants had very-high risk, with ∼10% had multiple ASCVD events and ∼50% had 1 ASCVD event and multiple high-risk conditions. This percentage is mildly greater than those from other studies in US, which reported a percentage of 43.0% to 58.0% ^9, 26, 27^. Among very-high-risk ASCVD, 17.0% to 27.0% had multiple ASCVD events ^9, 26-28^. One possible explanation between the mild disparity was that only patients with health insurance were included in other studies ^9, 26^. Moreover, our study remains the first nationally representative estimates of very-high-risk ASCVD while others were local. It’s notable that the percentage of participants with multiple ASCVD events or 1 ASCVD event and multiple high-risk conditions increased after 2007-2010, if CHD event was not restricted to within 1 year. This might be the result of persistently increased prevalence of diabetes ^4^, chronic kidney disease ^29^ and obesity ^1^, and decreased BP control after 2010 ^30^.

The relatively stable prevalence of ASCVD in the US differs from remarkable increase in some other countries such as China. Prevalent cases doubled and age-standardized prevalence of ASCVD increased significantly by 15% from 1990 to 2016 in China ^31^. The reasons are multifactorial, but could be explained at least in part, by the increased population growth and aging, prolonged life expectancy, concurrent declining cardiovascular mortality ^31^, and a low proportion of ideal cardiovascular health in these countries ^32^. CHD remains the most common type of ASCVD in US, doubles that of stroke. While in China, stroke is more common than CHD ^31^.

We observed substantial increase of lipid control from 1999-2002 to 2015-2018 in ASCVD adults, but the magnitude of increment decreased after 2006. Cardiovascular benefits of intensive lipid control in ASCVD patients have been evidenced ^33, 34^, followed by worldwide recommendations ^17, 18^. The ESC/EAS guideline recommends a non-HDL-C target of <85 mg/dL in very-high-risk ASCVD participants ^18^. In accordance with this recommendation, only 7.7% of participants achieved the target in US.

We showed that BP control declined after 2014 after a steady but slight increase from 1999-2002 to 2010-2014, such that a similar estimated proportion of adults had controlled BP in 2015-2018 as in 1999-2002. The trends of BP control in ASCVD adults were in line with that observed from general population with hypertension ^30^, but the control rate was higher in the ASCVD participants. These changes might be attributable to the updates of guideline recommendations on BP definition and control targets. In late 2013, the eighth Joint National Committee (JNC 8) published a report that recommended higher treatment BP threshold and goals for some adults ^35^ than JNC 7 ^36^. This shift toward less intensive treatment of hypertension might result in reduced BP control with current goals.

Although the proportion of ASCVD participants who achieved all 3 risk-factor control has been increasing, the absolute proportion was low (<20%). Recent declines in glycemic and BP control after 2010 to 2014, along with the worsening of other risk factors such as obesity ^1^, might portend a possible population-level increase in ASCVD-related morbidity and mortality. Indeed, global burden of CVD analyses revealed that age-standardized mortality from ischemic heart disease began to increase since 2014, in parts of the United States and United Kingdom ^12^. Although with 3-fold higher risk of ASCVD events,^9^ very-high-risk ASCVD adults who achieved risk-factor control decreased after 2014, while not-very-high-risk ASCVD increased. These findings emphasize urgent need to implement existing effective pharmacologic and lifestyle therapies to this large ASCVD population. The update of various guidelines with more rigorous risk-factor control targets ^7, 17^ should provide some optimism about the future of ASCVD in US.

Younger adults were significantly less likely to achieve lipid target, likely due to less statin therapy. Although the younger were more likely to achieve BP target (likely due to lower baseline BP), those with unmet target were receiving less BP-lowering treatments than the older. Younger adults with CHD had similar CV mortality ^37^, and those with stroke had an excess mortality than the older ^38^. The potential loss of lifetime productivity and greater lifetime financial burden in younger ASCVD adults ^39^ emphasize the need for early detection and management among these populations.

### Strengths and Limitations

This study has several strengths. NHANES is a large, nationally representative survey with a standardized protocol and rigorous quality control. Our analyses involved a large sample of adults with ASCVD collected from a continuous national survey. Trends in prevalence, control of risk factors and medication use in ASCVD were comprehensively analyzed over a 20-year period.

This study also has several limitations. First, analyses were based on self-reported ASCVD, which might cause misclassification or underestimation; however, it has been reported that self-reported coronary heart disease, myocardial infarction, and stroke is reliable ^40^. Second, ASCVD in our study did not include transient ischemic attack or peripheral artery disease, but this definition has also been widely used ^12-16^ and reflects CVD that causes major global mortality ^12^. Third, NHANES did not capture dosages of statins, therefore it’s unable to determine the trends of high-intensity statins use, one of the major topics in ASCVD. Fourth, we did not perform adjustment for multiple comparisons, potential type I error might exist.

## Conclusions

In this nationally representative survey of US adults from 1999 through 2018, the estimated prevalence of ASCVD generally remained stable, with over 60.0% had very-high risk. BP, lipid, and glycemic control decreased in very-high-risk ASCVD but increased in not-very-high-risk ASCVD after 2014.

## Data Availability

All original data are available at the NHANES website.

## Abbreviations

ACEI: angiotensin-converting enzyme inhibitor
ASCVD: atherosclerotic cardiovascular disease
ARB: angiotensin receptor blocker
BMI: body mass index
CCB: calcium channel blocker
CHD: coronary heart disease
DPP-4: dipeptidylpeptidase 4
FBG: fasting blood glucose
GLP-1: glucose like peptide-1
HDL-C: high-density lipoprotein cholesterol
LDL-C: low-density lipoprotein cholesterol
(LDL-C) NHANES: National Health and Nutrition Examination Surveys
SGLT2: sodium-glucose co-transporter-2
TC: total cholesterol

## Acknowledgements

We thank Jing Zhang from Shanghai Tongren Hospital for his help in data extraction from the NHANES database.

## Notes

**Funding:** This work was partly supported by Fundings for Clinical Trials from the Affiliated Drum Tower Hospital, Nanjing University School of Medicine (2022-LCYJ-PY-06 and 2022- YXZX-NFM-02).

**Conflict of interest:** None to declare.

### Competing Interest Statement

The authors have declared no competing interest.

### Funding Statement

This work was partly supported by Fundings for Clinical Trials from the Affiliated Drum Tower Hospital, Nanjing University School of Medicine (2022-LCYJ-PY-06 and 2022-YXZX-NFM-02)

### Author Declarations

The NHANES program was approved by the National Center for Health Statistics Ethics Review Board, and all participants provided written consent.

